# Topographic–prognostic gradients of cortical hypometabolism in temporal lobe epilepsy

**DOI:** 10.64898/2026.08.13.26360391

**Authors:** Jiajie Mo, Fatemeh Fadaie, Jack Lam, Donna Gift Cabalo, Jordan DeKraker, Alexander Ngo, Ke Xie, Ian Goodall–Halliwell, Daniel Mendelson, Ella Sahlas, Judy Chen, Rui Ding, Guan Zhou, Raul R. Cruces, Marlo Naish, Paul Bautin, Meaghan Smith, Youngeun Hwang, Raluca Pana, Jeff Hall, Olivier Aron, Aristides Hadjinicolaou, Roy Dudley, Sami Obaid, Alexander G. Weil, Zhong Zheng, Lin Sang, Qiang Guo, Yuguang Guan, Andrea Bernasconi, Neda Bernasconi, Kai Zhang, Boris C. Bernhardt

## Abstract

Anterior temporal lobectomy (ATL) remains the standard surgical treatment for pharmacoresistant temporal lobe epilepsy (TLE), yet long-term seizure freedom remains suboptimal. Neuroimaging studies show neocortical metabolic abnormalities beyond the mesiotemporal epicentre, but how such patterns inform resection extent remains unclear. We hypothesized that neocortical hypometabolism in TLE follows a quantifiable spatial gradient that can be translated into personalized surgical strategies. Our multicentre study included 358 participants across discovery, validation, and sensitivity analyses. Multimodal MRI and FDG-PET data were processed to derive vertex-wise structural, intensity, and metabolic features. Individual metabolic abnormalities were quantified using a normative asymmetry modelling approach. In the discovery cohort (227 patients undergoing ATL and 37 healthy controls), we characterized the topography of neocortical hypometabolism, and evaluated its correspondence to cytoarchitectural profiles, multimodal MRI features, and hippocampal measures. Three gradient-informed surgical metrics were evaluated in relation to seizure outcomes, with replication in an independent prospective validation cohort of 38 patients undergoing ATL. An additional sensitivity cohort comprising 56 surgical candidates, whose procedure spared the temporal neocortex was included to assess the robustness. Neocortical hypometabolism in TLE followed a spatially organized gradient, with the most severe hypometabolism at the hippocampal–neocortical interface that diminished with increasing geodesic distance (*r* = 0.955, *P*_perm_ < 0.001). Regions closer to the interface exhibited lower cytoarchitectonic differentiation and stronger FLAIR-related alterations. Hippocampal abnormalities also showed distance-dependent coupling with neocortical metabolism (*r* = 0.871, *P*_perm_ < 0.001). Among surgical metrics, greater resection of severe hypometabolism was associated with seizure freedom (OR = 1.448, *P* = 0.022). The association was replicated in the validation cohort. The present study identified a hypometabolic gradient in TLE, which covaries with cytoarchitectonic organization, microstructural changes, and hippocampal– neocortical interactions. The gradient provides a biologically grounded framework for precise surgical planning, emphasizing that targeting severe hypometabolism may optimize prognosis.

## Introduction

Anterior temporal lobectomy (ATL) remains an established surgical treatment for pharmacoresistant temporal lobe epilepsy (TLE), supported by randomized controlled trials and long-standing clinical observations.^1,2^ Despite substantial advances in neuroimaging acquisition, postprocessing techniques, presurgical evaluation, and surgical proficiency, long-term seizure freedom is not guaranteed. Indeed, approximately one third of patients continue to suffer from seizure recurrence despite surgical intervention.^2^ This persistent ceiling highlights a key clinical challenge in defining individualized resection boundaries, particularly along the lateral temporal cortex, where it is largely governed by a set and non-tailored resection extent.

Neuroimaging provides a unique opportunity to personalize surgical target definition beyond conventional anatomical boundaries.^3^ Complementing the utility of magnetic resonance imaging in localizing and lateralizing mesiotemporal pathology,^4–7^ [^18^F]fluorodeoxyglucose positron emission tomography (FDG-PET) can delineate atypical metabolism in TLE, with hypometabolism generally extending beyond mesiotemporal structures toward the temporal neocortex.^8^ Importantly, the extent of resection of PET hypometabolism has been shown to contribute to postoperative outcomes, suggesting that while “where” the presumed focus lies is important, “how much” abnormal cortex is removed may better facilitate individualized resection.^9^ However, a critical gap remains between imaging-derived abnormalities and surgical implementation, particularly in defining how far resections should extend beyond the mesiotemporal disease epicentre for a given patient. This question is supported by increasing evidence suggesting a spatially organized pattern of temporal hypometabolism in TLE, with abnormalities most pronounced in anterior regions and a progressive attenuation in the posterior direction. This continuous spatial variation suggests a gradient-like organization rather than a focal process.^10^ However, there is currently no quantitative approach to translate topographic patterns into discrete anatomical boundaries and to inform surgical strategies. Indeed, surgical extent is often determined heuristically rather than guided by quantitative imaging insights.

Here, we developed a neurocomputational framework (i) to quantify the topography of temporal neocortical hypometabolism gradient, (ii) to explore its potential biological substrates through analyses of cytoarchitectonic organization, multimodal alterations, and hippocampal–neocortical interactions, and (iii) to assess the utility of metabolism-informed resection metrics for surgical prognostication. This approach enables a quantitative translation of metabolic information into personalized resection prognostication (**Figure 1A–C**).

**Figure 1.**
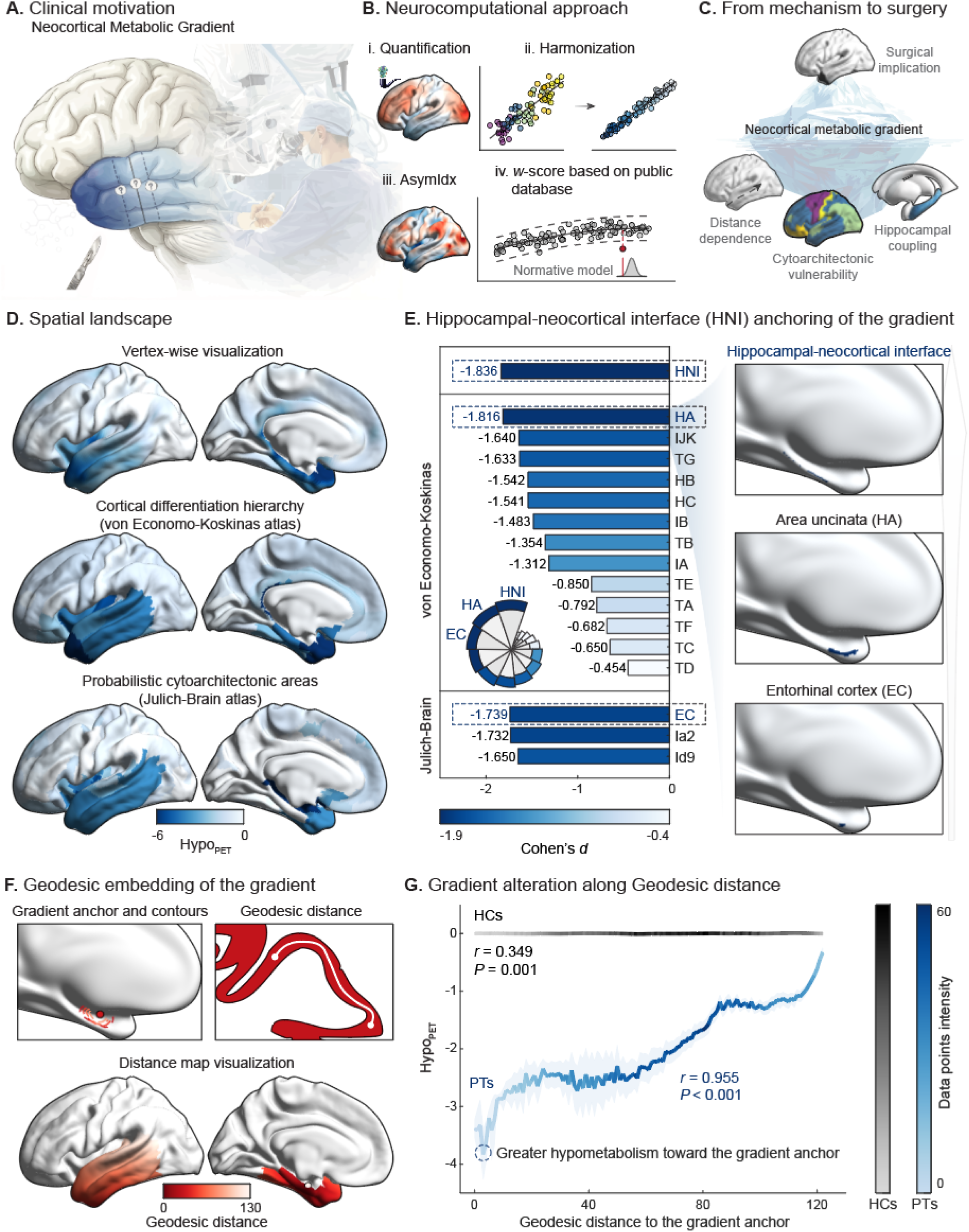
Neocortical metabolic gradients define a spatially organized epileptogenic continuum in TLE. **(A)** Schematic illustration showing spatially organized hypometabolism along the temporal neocortex, with greater abnormalities toward anterior regions; **(B)** Neurocomputational framework for gradient quantification. Multistep processing pipeline including feature quantification, harmonization, asymmetry index computation, and normative modeling to derive vertex-wise metabolic abnormality maps; **(C)** Conceptual framework linking hypometabolism to surgical targeting. The neocortical metabolic gradient is characterized by distance dependence, cytoarchitectonic vulnerability, and hippocampal coupling, providing a basis for gradient-informed surgical strategies; **(D)** Spatial distribution of neocortical metabolic abnormalities. Parcel-wise (Economo-Koskinas atlas and Julich-Brain atlas), and vertex-wise visualizations of Hypo_PET_ in patients, showing spatial hypometabolism gradient involving the temporal neocortex; **(E)** Identification of the gradient anchor. Bar and rose plots represented regional effect sizes (Cohen’s *d*) across temporal subregions, highlighting maximal abnormalities at the hippocampal-neocortical interface which was defined as the anchor of the metabolic gradient; **(F)** Geodesic embedding of the metabolic gradient. Schematic illustration of the gradient anchor and geodesic contours along the cortical surface, together with distance mapping projected onto the temporal neocortex. Color scale indicates increasing distance from the anchor; **(G)** Distance-dependent organization of metabolic abnormalities. The relationship between geodesic distance and Hypo_PET_ showed a strong monotonic pattern in patients (blue) but minimal spatial dependence in healthy controls (grey). Each solid line represents contour-wise mean values, shaded areas indicate 95% confidence intervals, and color intensity reflects data density. *Abbreviations*: Hypo_PET_ = normative modeling-derived *w*-score of [^18^F]fluorodeoxyglucose positron emission tomography asymmetry index; HA = Area uncinate; HB = Area parauncinata; HC = Area rhinalis limitans; TA = Area temporalis superior; TB = Area supratemporalis magnocellularis simplex; TC = Area supratemporalis granulosa; TD = supratemporalis intercalata; TE = Area temporalis propria; TF = Area fusiformis; TG = Area temporoporalis; FJK = Area frontoinsularis + Area piriformis frontalis; IA = Area insulae praecentralis; IB = Area insulae postcentralis; EC = Entorhinal cortex; Ia2 = Insular area 2; Ia9 = Insular area 9.

## Methods

### Study design and participants

We conducted a multicentre study comprising discovery, validation, and sensitivity cohorts of 358 participants. The discovery dataset consisted of 227 retrospective patients with pharmacoresistant TLE who underwent ATL at Beijing Tiantan Hospital and Beijing Fengtai Hospital recruited from 2018 to 2024, together with 37 healthy controls from the Centre d’exploration et de recherche multimodale et pluridisciplinaire (CERMEP) IDB-MRXFDG database for normative modelling to derive *w*-score measures.^11^ The validation cohort consisted of 38 patients undergoing ATL from an ongoing prospective study at Beijing Tiantan Hospital and Sanbo Brain Hospital (NCT06341075; from 2025). Sensitivity analyses were performed in an additional cohort of 56 patients who underwent selective amygdalohippocampectomy (SAH) or magnetic resonance-guided laser interstitial thermal therapy (MRgLITT). All patients underwent a multidisciplinary presurgical evaluation for epilepsy surgery (**Supplementary Method 1**). Detailed inclusion/exclusion criteria and the study flowchart were provided in **Supplementary Table 1** and **Supplementary Figure 1**.

The study was conducted in accordance with the Strengthening the Reporting of Observational Studies in Epidemiology (STROBE) guidelines^12^ and the Declaration of Helsinki. Ethical approval was obtained from the Institutional Review Board of Beijing Tiantan Hospital (KY2020-126-01). Written informed consent was obtained from all participants or their legal guardians.

### Imaging acquisition and preprocessing

All patients and healthy controls included in the analyses had complete multimodal imaging data, including T1-weighted, fluid attenuated inversion recovery (FLAIR) MRI, and FDG-PET, with acquisition protocols across centres described in **Supplementary Method 2**. Preprocessing was performed using a containerized implementation of *micapipe* (v0.2.0) within a Brain Imaging Data Structure (BIDS) framework.^13^ Cortical surface reconstruction was performed using *FastSurfer* (v2.0.0),^14^ incorporating native T1-weighted image reorientation, intensity non-uniformity correction (N4), and intensity normalization, followed by quality control and mask refinement.^15^ Intensity features were derived from T1-weighted and FLAIR images using T1w/FLAIR ratio mapping, including bias correction and white matter intensity normalization.^16^ Hippocampal anatomy was modelled using *HippUnfold* (v1.4.1), enabling topologically consistent surface representations in unfolded space.^17^ FDG-PET data were co-registered to native T1-weighted space, partial volume corrected, intensity-normalized to average pons uptake, and subsequently projected onto cortical and hippocampal surfaces.^18^ Neocortical features were mapped to fsLR-32k surface space, whereas hippocampal features were represented on *HippUnfold* surfaces; all features were subsequently smoothed along their respective surfaces using Gaussian kernels (10 mm full-width-at-half-maximum for the neocortex and 5 mm for the hippocampus).

For all patients, postoperative resection cavities were manually delineated on postoperative images by two experienced researchers (Dice coefficient = 0.803 ± 0.064). Cavity masks were registered to preoperative native space and projected onto cortical surfaces using nearest-neighbour volume-to-surface mapping to avoid interpolation-induced smoothing.^19^ All segmentations, registrations, and surface mappings were visually inspected to ensure accuracy. Full acquisition and preprocessing details are provided in the **Supplementary Method 3**.

### Feature extraction and quantification

Vertex-level multimodal features were extracted from the neocortex and hippocampus, including structural (thickness, curvature), intensity (midsurface FLAIR signal, T1w/FLAIR-derived myelin proxy), and metabolic (midsurface FDG-PET standardized uptake value ratio [SUVr]) metrics (**Supplementary Table 2**). To account for site-related variability, features were harmonized using *ComBat,* while preserving group, age, and sex effects.^20^ Hemispheres were aligned according to the epileptic focus, and asymmetry indices were computed between ipsilateral and contralateral regions. Individual deviations were quantified using normative modelling, with asymmetry features transformed into *w*-scores that also controlled for age and sex relative to asymmetries seen in healthy controls.^21^ Hypo_PET_, derived from the *w*-score of harmonized PET SUVr asymmetry, served as the main metric to characterize neocortical hypometabolism in subsequent analyses (**Supplementary Method 4** and **Supplementary Figure 2).**

### Gradient modelling and geodesic embedding

To quantify the spatial organization of neocortical hypometabolism, vertex-wise Hypo_PET_ maps were averaged to derive group-level patterns of abnormalities. Regional effects were evaluated across the Economo–Koskinas atlas^22^ (**Supplementary Figure 3**), Julich–Brain atlas^23^ and a hippocampal–neocortical interface (HNI) representation of the mesiotemporal transition zone. Geodesic distances were then computed along the cortical surface from the interface, and distance-dependent Hypo_PET_ profiles were quantified across successive contour bands to capture continuous variation in hypometabolism along the temporal neocortex.

### Cytoarchitectonic vulnerability

To assess microstructural vulnerability, cortical regions were stratified according to cytoarchitectonic differentiation, and complemented by layer-resolved cellular architecture derived from laminar organization, including layer thickness, cell size, and cell intensity (**Supplementary Figure 4A**). BigBrain-derived metrics, including principal cytoarchitectonic gradients (G1, G2), mean and skewness of intracortical staining profiles, were projected onto the surface.^24^ Distance-dependent changes in these features were evaluated along the geodesic axis to examine their relationship with metabolic abnormalities.

### Multimodal correspondence

To assess whether metabolic abnormalities are accompanied by structural alterations in TLE, we extracted the same set of structural, intensity, and metabolic features in patients and controls. We first assessed group-level differences across all features between patients and controls. We then quantified vertex-wise correlations between structural-intensity and metabolic features within each group separately. Finally, between-group differences in coupling strength were evaluated to identify features showing the most pronounced disease-related alterations (**Supplementary Method 5**).

### Hippocampal effects

We also examined associations between hippocampal features and neocortical Hypo_PET_ in patients. Multimodal hippocampal features were first evaluated using univariate analyses, followed by calculating a multivariate hippocampal abnormality index. Associations between hippocampal abnormalities and neocortical Hypo_PET_ were then quantified, and resulting effect sizes were examined along the geodesic axis (**Supplementary Method 6**).

### Surgical metrics and seizure outcome prediction

Three quantitative surgical metrics were derived: resection proportion of severe hypometabolism (proportion of cortex with Hypo_PET_ ≤ -1.96 standardized deviations resected), normalized resection volume, and resection boundary along the hypometabolic gradient (ratio of the 95th percentile Hypo_PET_ within the resection mask to that within the temporal lobe mask). Detailed definitions and computations are provided in **Supplementary Method 7**. Associations between these surgical metrics and seizure outcomes were assessed, with hippocampal asymmetry included for comparative purposes. Non-linear effects were explored using individual conditional expectation analyses,^25^ with restricted cubic splines used to identify potential inflection points. An independent validation cohort was used to confirm reproducibility of these surgical metrics and their association with outcomes. Seizure outcome was defined as seizure freedom (SF; Engel I) versus non-seizure freedom (NSF; Engel II–IV) at 1 year follow-up.^26^

### Statistical analysis

Group comparisons of imaging-derived measures were performed using two-sample *t* tests, with effect sizes quantified using Cohen’s *d*. Vertex-wise analyses were conducted within a general linear model framework, adjusting for age, sex, epilepsy duration, seizure frequency, and hippocampal volume using *SurfStat* Toolbox.^27^ Multiple comparisons were controlled using false discovery rate correction (*P*_FDR_ < 0.05). Spatial associations between imaging features and geodesic distance were assessed using Pearson correlation (*r*), while map-to-map spatial correlations were further validated using *BrainSMASH* permutation tests (10,000 permutations) to account for spatial autocorrelation.^28^ Associations between surgical metrics and seizure outcomes were evaluated using logistic regression, with odds ratio (OR) and 95% confidence intervals (CIs) reported. All statistical tests were two-sided, with significance set at *P* < 0.05. Analyses were performed in MATLAB R2024b (MathWorks, Natick, MA, USA).

## Results

### Cohort and clinical characteristics

A total of 358 participants were included, comprising 321 patients with TLE (mean age 29.1 ± 11.3 years; 153 females [47.7%]) and 37 healthy controls (mean age 38.0 ± 11.5 years; 20 females [54.1%]). Among patients, the mean epilepsy duration was 14.4 ± 9.8 years across cohorts. Postoperative seizure outcomes were available in all patients after a mean follow-up of 41.6 ± 22.4 months, with Engel I seizure freedom rates ranging from 74.2% to 88.9% across cohorts. Seizure frequency and clinical characteristics were comparable between discovery and validation cohorts, indicating overall consistency of patient profiles (**Table 1**).

**Table 1.** Demographic and clinical characteristics of the six study cohorts (n = 358)

### Neocortical metabolic gradients

Group comparisons revealed a distributed pattern of neocortical hypometabolism in patients, with the maximal metabolic deviation localized to the mesial temporal transition zone (**Figure 1D**). Although lateral temporal regions appeared visually involved, the strongest statistical effect converged at the HNI (*d* = -1.836, *P*_FDR_ < 0.001), corresponding to the mesiotemporal transition between hippocampal and neocortical territories (**Figure 1E**). Geodesic analyses demonstrated a marked distance-dependent organization of metabolic abnormalities away from the interface. In patients, Hypo_PET_ showed a strong monotonic relationship with geodesic distance, with hypometabolism progressively attenuating as distance from the HNI increased (*r* = 0.955, *P*_perm_ < 0.001), whereas the corresponding effect in controls was markedly weaker (*r* = 0.349, *P*_perm_ < 0.001). These findings indicate that neocortical hypometabolism in TLE follows a distance-dependent, spatially embedded gradient organization anchored at the HNI (**Figure 1F–G**).

### Cytoarchitectonic differentiation constrains metabolic vulnerability

This topographic metabolic gradient was also reflected in cortical cytoarchitecture, with effects decreasing from cortex with less laminar differentiation towards cortex with more marked lamination. The largest group differences were observed in agranular cortex (*d* = - 1.817, *P*_FDR_ < 0.001), whereas highly differentiated cortex showed smaller, yet still measurable effects (*d* = -0.484, *P*_FDR_ = 0.006) (**Figure 2A**). Multivariate layer-specific analyses revealed selective associations between laminar features and metabolic abnormalities, with layer II showing a significant relationship with Hypo_PET_ in patients (**Supplementary Figure 4B**). Consistently, regions proximal to the HNI were characterized by lower cytoarchitectonic differentiation. Histology-informed analyses further showed that BigBrain-derived gradients (G1, G2) and intracortical profile features (mean and skewness) varied systematically along the same geodesic axis and were significantly associated with Hypo_PET_ (**Figure 2B**). Together, these findings indicate that neocortical metabolic abnormalities are preferentially localized to less differentiated cortex and follow an intrinsic cytoarchitectonic axis of vulnerability.

**Figure 2.**
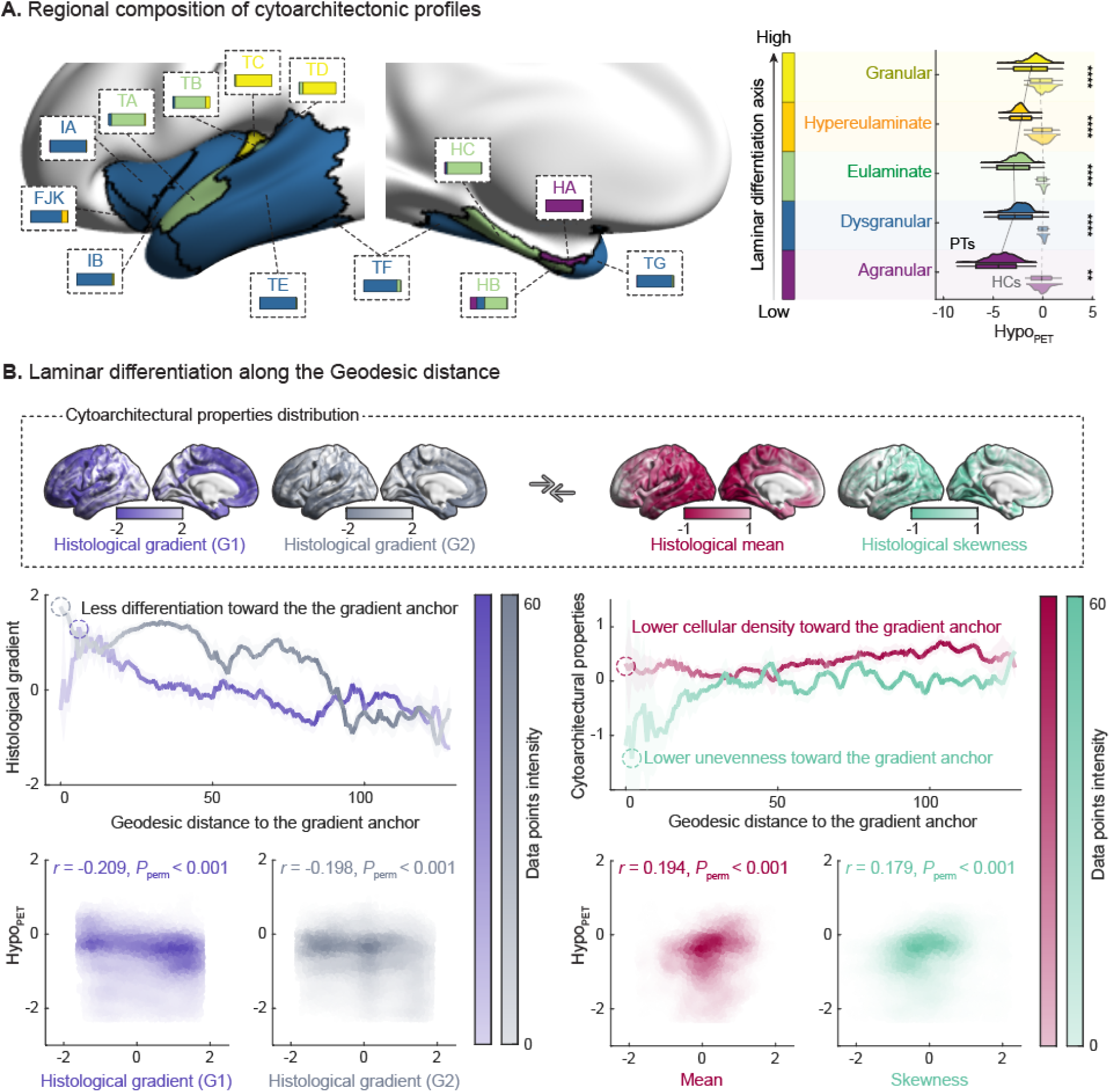
Cytoarchitectonic differentiation reflects metabolic vulnerability. **(A)** Cortical regions were classified according to cytoarchitectonic differentiation, spanning a gradient from less differentiated (agranular, purple) to highly differentiated (granular, yellow) cortex. Surface maps show the spatial distribution of cytoarchitectonic classes across the temporal lobe, alongside representative laminar differentiation axes. Stacked bar plots depict the relative proportion of each class within regions, with colors indicating their position along the differentiation gradient. Violin plots illustrate group differences in Hypo_PET_ across cytoarchitectonic classes in patients (PTs) and healthy controls (HCs), demonstrating progressively greater hypometabolism toward less differentiated cortex; **(B)** Cytoarchitectonic and histological organization along the geodesic axis. Top: surface projections of BigBrain-derived cytoarchitectonic features, including principal histological gradients (G1, G2) and intracortical profile metrics (mean and skewness). Middle: distance-dependent variation of these features along the geodesic axis from the gradient anchor, illustrating systematic changes in laminar differentiation and cellular architecture. Solid lines represent contour-wise mean values, shaded areas indicate 95% confidence intervals, and color intensity reflects data density. Bottom: spatial associations between cytoarchitectonic features and Hypo_PET_. Density plots show vertex-wise relationships, with corresponding correlation coefficients and permutation-based significance. *Abbreviations*: Hypo_PET_ = normative modeling-derived *w*-score of [^18^F]fluorodeoxyglucose positron emission tomography asymmetry index; HA = Area uncinate; HB = Area parauncinata; HC = Area rhinalis limitans; TA = Area temporalis superior; TB = Area supratemporalis magnocellularis simplex; TC = Area supratemporalis granulosa; TD = supratemporalis intercalata; TE = Area temporalis propria; TF = Area fusiformis; TG = Area temporoporalis; FJK = Area frontoinsularis + Area piriformis frontalis; IA = Area insulae praecentralis; IB = Area insulae postcentralis.

### Multimodal correspondence reveals dominant FLAIR-metabolic coupling alterations

Group comparisons across multimodal features revealed distinct alteration patterns in patients relative to controls. Structural features such as cortical thickness and curvature showed moderate reductions, whereas intensity-based features demonstrated divergent changes, with midsurface FLAIR intensity exhibiting the largest increases. Moreover, metabolic measures showed the most pronounced decreases, indicating marked hypometabolism (**Figure 3A**). Among all features, midsurface FLAIR intensity showed the strongest association with Hypo_PET_ in patients (*r* = -0.413, *P*_perm_ < 0.001), whereas other features such as cortical thickness (*r* = 0.125, *P*_perm_ < 0.001), curvature (*r* = 0.085, *P*_perm_ < 0.001) and myelin proxy (*r* = 0.125, *P*_perm_ < 0.001) exhibited weaker relationships. The magnitude of change in coupling between patients and controls was also greatest for midsurface FLAIR intensity (Δ*r* = 0.420), indicating a disease-related reorganization of structure-metabolism coupling (**Figure 3B**). These findings suggest that hypometabolism in TLE is most closely associated with FLAIR-related signal alterations, potentially reflecting gliosis-related tissue changes.

**Figure 3.**
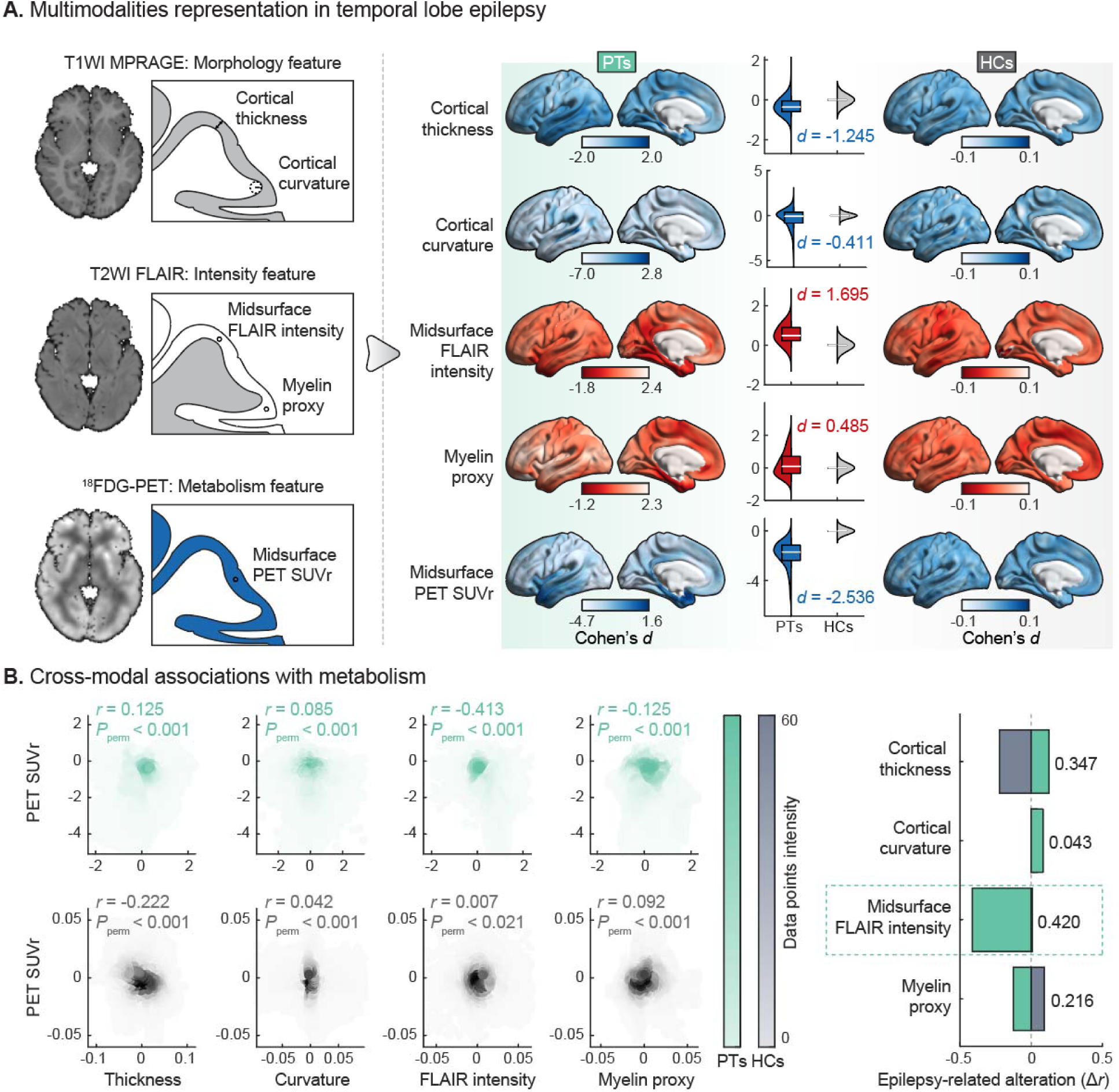
Correspondence to MRI alterations. **(A)** Correlation to structural and intensity features. Representative features derived from T1-weighted MRI (cortical thickness and curvature), FLAIR imaging (midsurface FLAIR intensity and T1/FLAIR-derived myelin proxy), and FDG-PET (midsurface PET standardized uptake value ratio [SUVr]) are shown. Surface maps illustrate group-level differences in distribution between patients (PTs) and healthy controls (HCs). Accompanying violin plots depict the distribution of feature values across vertices, while overlaid box plots indicate median and interquartile range. Structural features exhibit moderate reductions, whereas intensity-based features show divergent alterations, with midsurface FLAIR intensity demonstrating the largest increases. In contrast, metabolic measures show the most pronounced decreases, indicating widespread hypometabolism; **(B)** Cross-modal associations with metabolic abnormalities. Vertex-wise correlations between structural-intensity and metabolic features are shown for PTs (green) and HCs (grey), with corresponding permutation-based significance. Density plots illustrate spatial correspondence across the cortex. Among all features, midsurface FLAIR intensity exhibits the strongest association with metabolic abnormalities in PTs. Bar plots summarize differences in coupling strength between groups (Δ*r*), highlighting midsurface FLAIR intensity as showing the largest disease-related effect. *Abbreviations*: MPRAGE = magnetization-prepared rapid acquisition gradient echo; FLAIR = fluid-attenuated inversion recovery; [^18^F]FDG-PET = [^18^F]fluorodeoxyglucose positron emission tomography; SUVr = standardized uptake value ratio; PTs = patients; HCs = healthy controls.

### Hippocampal effects along the neocortical metabolic gradient

Ipsilateral hippocampal features showed distinct value distributions and spatially heterogeneous patterns across the hippocampal surface. Univariate analyses revealed that associations between hippocampal features and neocortical Hypo_PET_ were concentrated in the mesiotemporal region near the HNI. Among individual features, hippocampal Hypo_PET_ (|*d*| = 2.742) showed the strongest effects, followed by thickness (|*d*| = 2.030) and FLAIR intensity (|*d*| = 0.864). Multivariate analysis using hippocampal Mahalanobis distance yielded a consistent spatial pattern (**Figure 4A**). In addition to the distance-dependent pattern observed for metabolic abnormalities, hippocampal covariance effects exhibited a similar geodesic organization, with feature-specific associations varying systematically along the distance-based axis. Notably, hippocampal Mahalanobis distance, a multivariate abnormality index integrating all hippocampal structural-intensity-metabolic features, demonstrated a significant positive association with geodesic distance (*r* = 0.871, *P*_perm_ < 0.001), indicating progressively increased coupling toward the gradient origin (**Figure 4B**). These findings indicate that hippocampal abnormalities are coupled to neocortical metabolic dysfunction in a distance-dependent manner, supporting a gradient-aligned hippocampal–neocortical interaction.

**Figure 4.**
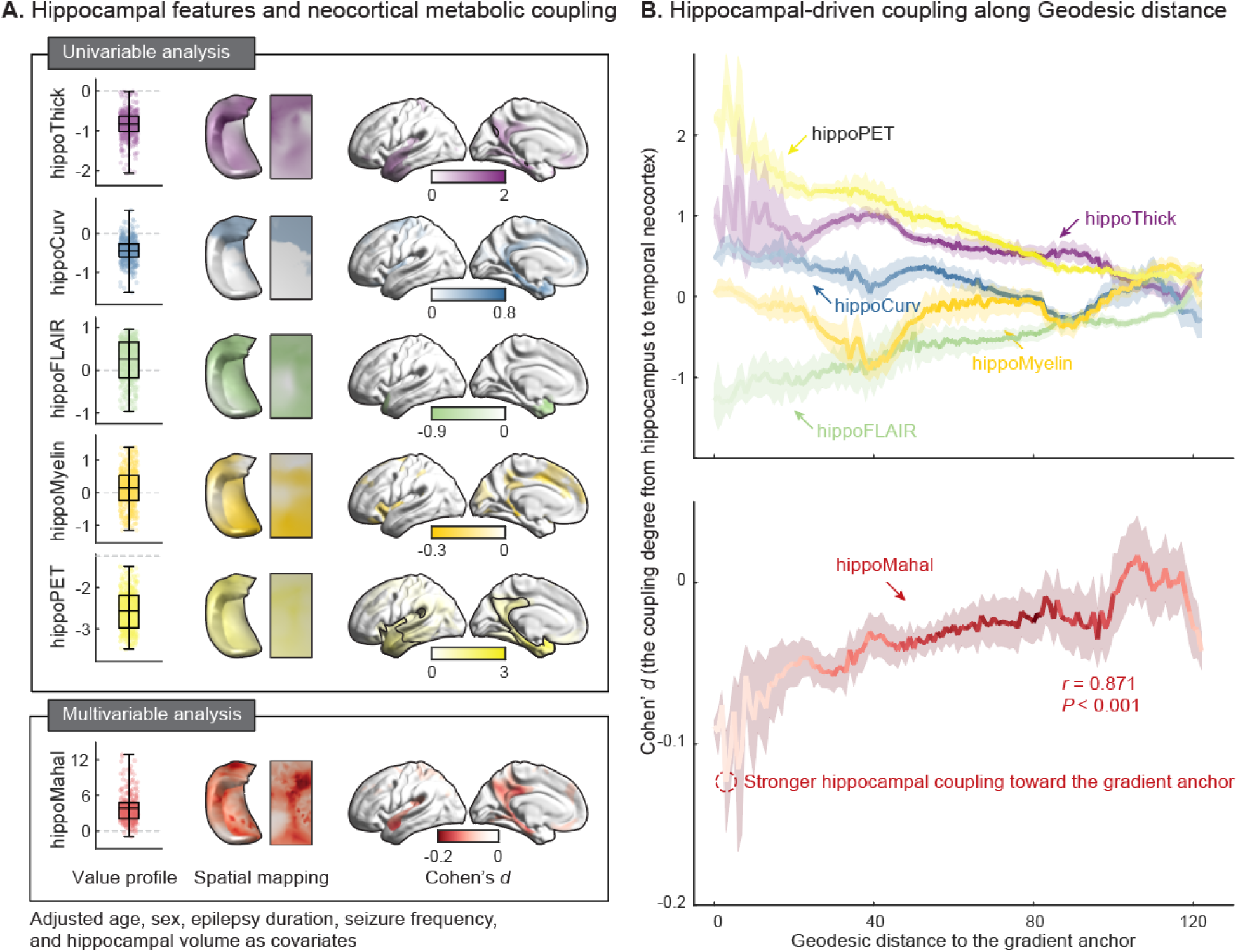
Associations to hippocampal findings. **(A)** Hippocampal features and their neocortical metabolic associations. Boxplots illustrate the distribution of ipsilateral hippocampal feature values across patients, with overlaid scatter points representing individual observations. Spatial representations include hippocampal surface maps in native and unfolded space, illustrating feature distributions across the hippocampus. Surface maps display the associations between hippocampal features and neocortical Hypo_PET_, presented for both univariate analyses of individual features and multivariate analysis using hippocampal Mahalanobis distance. All models were adjusted for age, sex, epilepsy duration, seizure frequency, and hippocampal volume. Color scales represent effect sizes (Cohen’s *d*), and outlined regions indicate statistical significance after false discovery rate correction; **(B)** Distance-dependent organization of hippocampus-driven coupling. Associations between hippocampal features and neocortical metabolism are plotted along the geodesic distance to the gradient anchor. Feature-specific coupling profiles show systematic variation along the gradient axis. The multivariate hippocampal Mahalanobis distance increased progressively with geodesic distance, indicating a distance-dependent organization. As hippocampal Mahalanobis distance was inversely related to neocortical Hypo_PET_, lower values reflect stronger hippocampal-neocortical coupling. Solid lines represent contour-wise mean values, shaded areas indicate 95% confidence intervals, and color intensity reflects data density. *Abbreviations*: Hypo_PET_ = normative modeling-derived *w*-score of [^18^F]fluorodeoxyglucose positron emission tomography asymmetry index; hippoThick = hippocampal thickness; hippoCurv = hippocampal curvature; hippoFLAIR = hippocampal FLAIR intensity; hippoMyelin = hippocampal T1w/FLAIR-derived myelin proxy; hippoPET = hippocampal PET standardized uptake value ratio; hippoMahal = hippocampal Mahalanobis distance.

### Gradient-informed surgical metrics relate to postoperative seizure outcome

Vertex-wise comparisons revealed modest but significant differences in neocortical metabolic patterns between SF and NSF patients (*d* = -0.349, *P* = 0.028), whereas resection cavity distributions showed no significant group differences (*P* = 0.144) (**Figure 5A**). Cohort composition and surface-based illustration of the three surgical metrics related to seizure outcomes were shown in **Figure 5B** and **Figure 5C**. Among them, only the resection proportion of severe hypometabolism was significantly associated with SF (OR = 1.448, 95% CI = 1.056–1.986; *P* = 0.022), whereas clinical information (**Supplementary Figure 5**), hippocampal volume (**Supplementary Figure 6**), normalized resection volume and resection boundary along the hypometabolic gradient showed no significant association with outcomes. This association remained robust after controlling for ipsilateral extra-temporal hypometabolism (**Supplementary Figure 7**). ICE modelling revealed a monotonic increase in the probability of SF with greater hypometabolic core. RCS analysis further identified a significant inflection point at 53.38% (*P* = 0.038), above which the probability of seizure freedom increased more steeply with increasing resection proportion of severe hypometabolism (**Figure 5D**), although the marginal prognostic benefit appeared to diminish beyond approximately 76.25% resection (**Supplementary Figure 8**). These findings were reproduced in an independent validation cohort, which demonstrated a consistent trend (**Figure 5E**). Illustrative cases further highlight this relationship: a patient with greater resection proportion of severe hypometabolism achieved SF, whereas a patient with more diffuse abnormalities and limited resection proportion showed persistent seizures (**Figure 5F**). Additional analyses in the sensitivity cohort demonstrated that patients undergoing ATL exhibited significantly more severe neocortical hypometabolism than those treated with SAH/MRgLITT, particularly within anterior and mesial temporal regions (peak Cohen’s *d* = - 1.707, *P*_FDR_ < 0.05; **Supplementary Figure 9A**). In contrast, no significant differences in neocortical metabolic patterns were observed between SF and NSF patients within the sensitivity cohort (**Supplementary Figure 9B**). Collectively, these results demonstrate that surgical engagement of the resection proportion of severe hypometabolism is a promising indicator of favourable seizure outcomes.

**Figure 5.**
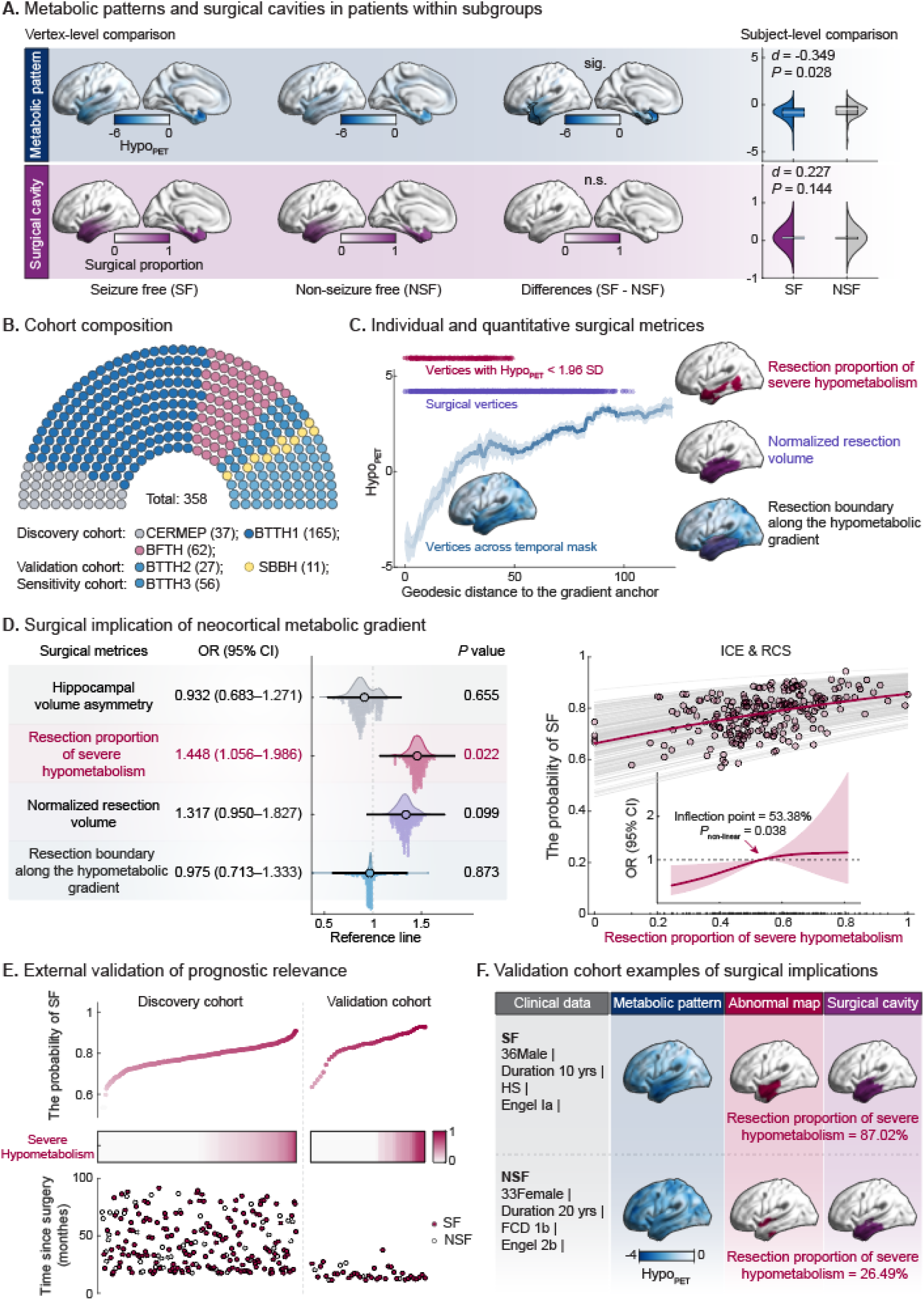
Associations between cortical hypometabolism, resections, and seizure outcome. **(A)** Spatial differentiation of metabolic pattern and surgical cavities between outcome groups. Vertex-wise comparisons of neocortical metabolic patterns and surgical cavities between seizure-free (SF) and non-seizure-free (NSF) patients show no significant group differences. Surface maps illustrate group-average patterns, and violin plots depict subject-level distributions with embedded box plots indicating median and interquartile range; **(B)** A parliament plot displays the distribution across discovery and independent validation cohorts; **(C)** Definition and spatial representation of gradient-informed surgical metrics. Vertex-wise profiles along the geodesic axis show Hypo_PET_ (blue), resected vertices (purple), and metabolically abnormal vertices defined by Hypo_PET_ ≤ -1.96 standard deviations (red). These components define the relationship between metabolic abnormality and surgical resection, from which three quantitative metrics were derived: resection proportion of severe hypometabolism, normalized resection volume, and resection boundary along the hypometabolic gradient. Surface-based illustrations depict the spatial representation of each metric; **(D)** Logistic regression models show that only the resection proportion of severe hypometabolism is significantly associated with SF, whereas the remaining features are not. Forest plots display odds ratios (OR) with 95% confidence intervals (95% CI), and violin plots illustrate the distribution of values across subjects. Individual conditional expectation (ICE) curves demonstrate a monotonic increase in the probability of SF with greater resection proportion of the severe hypometabolism. Restricted cubic spline (RCS) analysis identifies an inflection point (53.38%), above which the probability of SF increases more steeply; **(E)** Replication of the relationship between resection proportion of severe hypometabolism and probability of SF in an independent, prospective validation cohort, showing consistent positive trends. Scatter plots illustrate individual-level distributions, and color gradients indicate resection proportion of severe hypometabolism. The time since surgery for each patient was also displayed; **(F)** Representative validation cohort cases illustrating gradient-informed surgical implications. The first patient, with greater resection proportion of severe hypometabolism, achieved seizure freedom, whereas the second patient, with more diffuse abnormalities and lower resection proportion, showed persistent seizures. Panels display clinical information, metabolic patterns, abnormality maps, and surgical cavities. *Abbreviations*: Hypo_PET_ = normative modeling-derived *w*-score of [^18^F]fluorodeoxyglucose positron emission tomography asymmetry index; SF = seizure-free; NSF = non-seizure-free; CERMEP = Centre d’exploration et de recherche multimodale et pluridisciplinaire; BTTH = Beijing Tiantan Hospital; BFTH = Beijing Fengtai Hospital; SBBH = Sanbo Brain Hospital; SD = standard deviation; OR = odds ratios; 95% CI = 95% confidence intervals; ICE = individual conditional expectation; RCS = restricted cubic spline; yrs = years; FCD = focal cortical dysplasia; HS = hippocampal sclerosis.

## Discussion

The current study quantified topographic patterns of temporal neocortical hypometabolism in TLE and clarified their associations with surgical outcomes. We identified a gradient of hypometabolism that progressively decayed with increasing distance from the hippocampal– neocortical transition area, and which co-varied with cytoarchitectonic differentiation, FLAIR alterations, and measures of hippocampal pathology. As included patients all underwent epilepsy surgery subsequent to neuroimaging, we additionally examined associations with postoperative outcome and found that more complete resection of severe hypometabolism was associated with improved seizure freedom.^9^ The latter findings were not explainable by total resection volume, and persisted when controlling for extra-temporal hypometabolism or hippocampal volume abnormalities, which are otherwise recognized factors contributing to postsurgical seizure outcome. These findings support the potential utility of gradient-informed assessment of cortical hypometabolism for both the characterization and presurgical evaluation in TLE.

Our findings provide new evidence for topographic, neurobiological, and pathological substrates of temporal neocortical metabolism in TLE. First, we observed maximal effects near the hippocampal–neocortical transition area. Structural models of cortical organization suggest that allocortical, periallocortical, and paralimbic regions differ from more clearly laminated isocortex in both architecture and connectivity.^29^ Recent large-scale work further suggests that cortical differentiation follows a continuous organizational axis extending from allocortical/periallocortical regions toward primary sensory cortex. Within this framework, less differentiated cortex may occupy a biologically distinct architectonic position characterized by greater plasticity and reduced inhibitory stabilization, which may increase susceptibility to recruitment into epileptic networks and prolonged hyperexcitability, ultimately contributing to interictal hypometabolism.^30^ These findings are also closely related to our findings on differential hypometabolism across different cytoarchitectonic territories, as suggested by parallel contextualization of BigBrain 3D histology data.^24^ Second, the close association between TLE-related FLAIR signal increases and PET-derived hypometabolism potentially reflects gliosis-associated remodelling of the metabolic microenvironment. Histopathological and multimodal imaging studies have linked increased FLAIR signal in TLE to astroglial activation, glial fibrillary acidic protein-related gliosis,^31,32^ and preferential microstructural disruption within limbic and paralimbic cortices across ipsilateral temporolimbic regions.^33–35^ Such alterations may disrupt coordinated metabolic coupling between neurons and glial cells and glucose utilization, potentially contributing to reduced [^18^F]FDG uptake despite preserved or even increased cellular density.^36^ Third, the observed hippocampal–neocortical coupling supports a distributed network interpretation of neocortical hypometabolism in TLE. Connectome analyses in TLE have identified the hippocampus and paralimbic cortex as key epicentres within temporolimbic networks, from which structural and functional abnormalities preferentially extend along anatomical connectivity pathways.^37^ In the current study, multiple hippocampal features demonstrated distance-dependent coupling with neocortical hypometabolism along the geodesic axis, while the multivariate hippocampal abnormality index showed an even stronger and more spatially consistent relationship. Together, these observations support close interactions between hippocampal pathology and distributed metabolic abnormalities across interconnected temporolimbic regions, where propagation along anatomical connectivity pathways may contribute to the continuous, distance-dependent gradient-like pattern observed in the present study.

There have been substantial advances in neuroimaging technology and protocols, which have led to improved identification of epileptic pathology and surgical target definition.^38^ However, the outcome gains after ATL have remained modest, with SF rates showing little improvement since early randomized trials^1^ and substantial variability across centres (40%– 92.9%).^2^ Accumulating evidence suggests that case presentation may change over time, with changes in risk factors and imaging findings in TLE cases referred to tertiary centres.^3,38^ In the current study, more complete resection of severe hypometabolism was associated with improved seizure freedom, whereas overall surgical extent and other surgical metrics were not significantly associated with outcome. The key finding indicates that individualized metabolic abnormalities may provide clinically relevant information beyond conventional anatomical resection measures. At the same time, postoperative outcome in TLE is likely governed by multiple additional factors. First, the burden and spatial extent of hippocampal pathology remain central to current models of TLE, supported by recent studies demonstrating that hippocampal abnormalities and their extent of resection remain closely related to postoperative seizure outcomes.^19^ Second, pathological and clinical heterogeneity may also contribute to outcome variability. In the present cohort, patients with malformations of cortical development tended to show less favourable outcomes compared with pure hippocampal sclerosis, although these differences did not reach statistical significance, suggesting that individualized imaging-informed surgical metrics may capture patient-specific disease burden beyond conventional clinical categories. Third, abnormalities extending beyond the temporal lobe may also be relevant. The concept of “temporal-plus” epilepsy indicates that seizure-generating networks can involve adjacent regions and has been associated with substantially higher risk of surgery failure.^39^ In this context, our findings support the view that neocortical abnormalities in TLE may extend along distributed temporolimbic networks rather than remaining confined to mesial temporal structures alone. Notably, patients undergoing SAH/MRgLITT in our sensitivity cohort still achieved favourable seizure outcomes (78.2%) despite relative sparing of the temporal neocortex. Compared with ATL cases, these patients exhibited substantially less severe neocortical hypometabolism, suggesting that extensive neocortical resection may not be necessary when metabolic abnormalities remain more limited. Viewed from another perspective, this observation further supports the notion that severe neocortical hypometabolism identifies biologically relevant epileptic tissue, and that engagement of these metabolically abnormal regions, rather than resection extent *per se*, may represent a critical determinant of postoperative seizure control. Together, these findings support a shift from anatomy-based to biology-informed surgical planning in TLE.

This study has several limitations. First, our analyses were based on preoperative FDG-PET, and future studies incorporating postoperative metabolic imaging are needed to determine whether gradient-informed surgical targeting improves long-term seizure outcomes. It will also clarify how residual or reorganized hypometabolic gradients relate to postoperative network remodelling. Second, although the present work focused on TLE, the association between resection of severe hypometabolism and seizure outcome suggests a potential direction for future studies in other epilepsies as well as surgical contexts. Such work will be needed to determine whether hypometabolism-informed metrics may serve as robust and generalizable imaging biomarkers for surgical prognosis.

In conclusion, we quantitatively define, biologically contextualize, and clinically translate a neocortical metabolic gradient in TLE, thereby informing clinical decision making, supporting individualized resection strategies, and ultimately improving patient outcomes.

## Data Availability

All data produced in the present study are available upon reasonable request to the authors

## Data availability

Pseudonymized data will be shared by request from a qualified academic investigator for the sole purpose of replicating procedures and results presented in the article, and as long as data transfer is in agreement with general data protection regulation and decisions by the Ethical Review Board of Beijing Tiantan Hospital, Beijing Fengtai Hospital and Sanbo Brain Hospital, which should be regulated in a material transfer agreement.

## Acknowledgments

Not available.

